# The role of mouthwash sampling in SARS-CoV-2 diagnosis

**DOI:** 10.1101/2021.07.22.21260760

**Authors:** Asaf Biber, Dana Lev, Michal Mandelboim, Yaniv Lustig, Geva Harmelin, Amit Shaham, Oran Erster, Eli Schwartz

## Abstract

**Background:** The current practice of COVID-19 diagnosis worldwide is the use of oro-nasopharyngeal (ONP) swabs. Our study aim was to explore mouthwash (MW) as an alternative diagnostic method, in light of the disadvantages of ONP swabs.

**Methods:** Covid-19 outpatients molecular-confirmed by ONP-swab were repeatedly examined with ONP-swab and MW with normal-saline (0.9%). Other types of fluids were compared to normal-saline. The Cq values obtained with each method were compared.

**Results:** Among 137 pairs of ONP-swabs and MW samples, **84.6%** (116/137) of ONP-swabs were positive by at least one of the genes (N, E, R). However MW detected 70.8% (97/137) of samples as positive, which means **83.6%** (97/116) out of positive ONP-swabs, missing mainly Cq value>30. In both methods, the N gene was the most sensitive one. Therefore MW samples targeting N-gene, which was positive in 95/137 (69.3%), is comparable to ONP-swabs targeting E and R genes which gave equal results – 95/137 (69.3%) and 90/137 (65.7%) respectively.

Comparing saline MW to distilled-water gave equal results, while commercial mouth-rinsing solutions were less sensitive.

**Conclusions:** MW with normal-saline, especially when tested by N gene, can effectively detect COVID-19 patients. Furthermore, this method was not inferior when compared to R and E genes of ONP-swabs, which are common targets in many laboratories around the world.

## Introduction

The COVID-19 pandemic has prompted an unprecedented global consumption of diagnostic equipment, and the demand for diagnostic tests continues to rise. As of September 4, 2020, the WHO has shipped 9,826,519 swabs worldwide to insure supply for low-middle-income countries.^1^ In light of the tremendous need to increase the availability of diagnostic tests, the NIH launched the Rapid Acceleration of Diagnostics (RADx) program, to support the development, production and deployment of rapid accurate tests.^2^

SARS-CoV2, the etiological agent of the ongoing COVID-19 pandemic, is transmitted through respiratory droplets. Patients with COVID-19 have demonstrated high viral loads in their upper and lower respiratory tracts beginning as early as 5-6 days before the onset of symptoms.^3-6^ The current methods of diagnosis of SARS-CoV2 include detection of the virus by genomic techniques using reverse transcription polymerase chain reaction (RT-PCR).^7^

Oro- or nasopharyngeal sampling is currently the gold standard for diagnosis, with a range of sensitivity results, presumably due to different collection methods and laboratory techniques.

The major disadvantages of nasopharyngeal swabs are the necessity of swabs and medium, which are largely unavailable in many parts of the world, their cost, the need to train workers to collect the specimens, as well as discomfort to the patient and the potential risk of infection for the examiner. These disadvantages motivate efforts to explore different sampling methods.

One proposed method to replace the use of swabs was the collection of saliva. This method was tested in several recent studies^8^, with contradicting conclusions when compared to nasopharyngeal swabs. For example, in a clinical trial conducted in Canada, which compared 91 pairs of nasopharyngeal swabs and saliva samples, sensitivity was 89% for nasopharyngeal swabs and significantly less for saliva - only 72%.^9^ The difference in sensitivity was greatest for sample pairs collected later during the illness course.^9^ On the contrary; the same comparison was made in Connecticut, USA, for 70 patients and yielded a higher percentage of positive saliva samples than nasopharyngeal swab samples up to 10 days after the diagnosis. In addition, more copies of SARS-CoV-2 RNA were detected in the saliva specimens.^10^ Nonetheless, in August 2020 the FDA issued an emergency use authorization (EUA) for a self-collecting saliva kit, named SalivaDirect - a RT-qPCR test from saliva collected by healthcare providers, which uses Proteinase K and heating to extract RNA.^11^

Herein we present the utility of MW samples for the detection of SARS-CoV-2, a method that has not been thoroughly examined. In contrast to ONP swabs, MW is a noninvasive, simple and inexpensive test that can be easily performed by the patient him or herself. This study examined a variety of fluids for mouth rinsing.

## Methods

### Study design

This was a community-based, prospective trial, to evaluate the effectiveness of MW in the detection of SARS-CoV-2 as a diagnostic method. The study was conducted mainly in non-hospital facilities dedicated for COVID-19 patients in isolation. Patients were followed during their stay in these facilities.

Ethical approval was obtained from Sheba Medical Center and informed consent was obtained from all patients. The trial was done in accordance with the principles of the declaration of Helsinki.

### Study population

The study population included adult men and non-pregnant women (>18 years old), with molecular confirmation of COVID-19 by RT-PCR. All patients were either asymptomatic or with mild symptoms.

#### Exclusion criteria

age under 18 years. Patients with severe infection (defined as need for invasive or non-invasive ventilator support, ECMO or shock requiring vasopressor support) were not included.

### Sample collection methodology

Between July and September 2020, we collected 361 samples from a total of 96 confirmed COVID-19 outpatients: 137 ONP swabs, 137 saline MW samples, 59 distilled water wash, 12 commercial MW solution containing alcohol and 16 commercial MW solution without alcohol.

Patients were repeatedly examined both for MW and ONP swabs. Due to the fact that the sensitivity of the test is affected by the examiner who performs the swab, and differences between labs, the same medical staff performed the swab sampling during the entire trial. All RT-qPCR tests were conducted by the Israel Central Virology Laboratory, using the same protocol, as detailed below.

#### Oro-Nasopharyngeal swabs procedure

The sampling guidelines in Israel instruct to insert first a swab into the posterior pharynx and tonsillar areas, followed by inserting a flexible swab through the nostril to the nasopharynx. The two swabs are placed together in a single tube to maximize test sensitivity.^12^ Therefore, the standard policy in Israel is actually an ONP swab testing.

#### Mouthwash sampling

Patients were asked to rinse and gargle 10 cc of normal saline (0.9%) for about 10-20 seconds and then spit the fluid into a sterile container.

In addition, other solutions were tested with a small number of patients; in order to compare it to the standard saline solution, which included:

a. Distilled water
b. Commercial mouth wash containing alcohol (Listrine®, manufactured by Johnson & Johnson)
c. Commercial mouth wash without alcohol (Orbitol®, manufactured by COSMOPHARM LTD.)

All samples were examined by RT-qPCR, as described below.

### Nucleic acid extraction

Samples were inactivated upon arrival to the laboratory, by heating at 70°C for 30 minutes. Subsequently, 400 µl were taken from each sample; mouth wash or ONP swab medium, and then the total nucleic acid content was extracted using MagLead 12gC (Precision System Science Co. Ltd, Japan) in 50 µl elusion buffer.

#### Real Time PCR

The presence of the viral RNA was detected using the Seegene Allplex CoV19 detection kit, according to the manufacturer’s instructions (http://www.seegene.com/assays/allplex_2019_ncov_assay). Briefly, the test detects three viral genes: envelop (E), nucleocapsid (N) and RNA-dependent RNA polymerase (RdRp). The integrity of the extraction procedure is monitored using an internal control that is inserted into the sample prior to the extraction procedure. The PCR integrity is monitored using a CoV19 positive control. Following mix assembly, the samples were analyzed using the Bio-Rad CFX96 thermal cycler, and its accompanying software, CFX Maestro (https://www.bio-rad.com/).

### Statistical analysis

Comparison between the Cq values obtained for the different reactions performed in the PCR test (E,N,R) for each method (ONP swab/MW), was evaluated by paired samples t-test. A *P* value < .05 was considered as statistically significant.

## Results

During a period of two months, 137 pairs of samples of MW and ONP swabs were collected. The study included 96 outpatients - 38 females and 58 males between ages 18-73. Among them, 20.5% were asymptomatic, while 79.5% presented with mild symptoms of fever, headache, malaise, cough, myalgia and more.

### Performance of ONP swabs

Among 137 tests there were **84.6%** positive cases for at least one of the genes. A Cq value of ≤ 40 was considered as a positive result. Among the 3 tested targets (N, E, R), the N gene reaction was the most sensitive. The N gene was positive in **114** tests, compared to **95** and **90** in the E and R genes respectively. It should be noted that with the exception of two weakly-positive tests (Cq= 38, 39), whenever R or E gene was positive, the N gene was positive as well (Figure 1a).

**Figure 1a:**
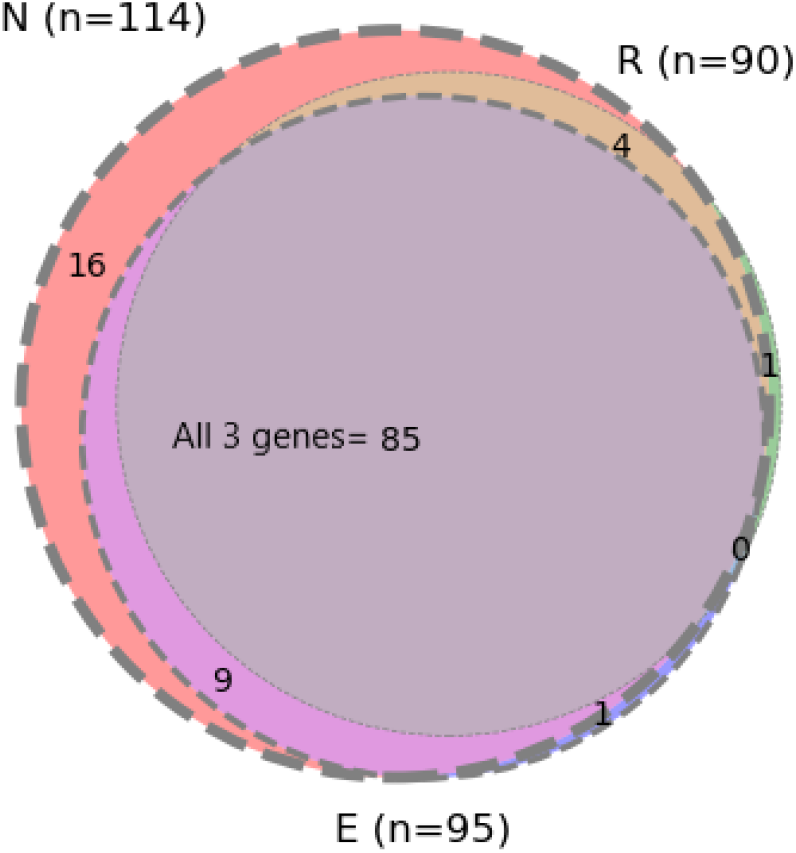
Number of positive results of **Oro-nasopharyngeal swab (N=137 tests)** in each target (N, E, R genes) and their overlapping.

### Performance of saline MW solution

Among 137 tests there were **70.8%** positive cases by at least one of the genes. Among the 3 tested targets (N, E, R), the N gene reaction seems to be the most sensitive. The N gene reaction was positive in **95** tests, compared to **69** in both E and R gene reactions. Similarly to the ONP swabs, the N gene reaction was almost always positive when R or E reactions were positive, with exception of two weak-positive tests, with high Cq values (39,40) and negative N gene target (Figure 1b).

**Figure 1b:**
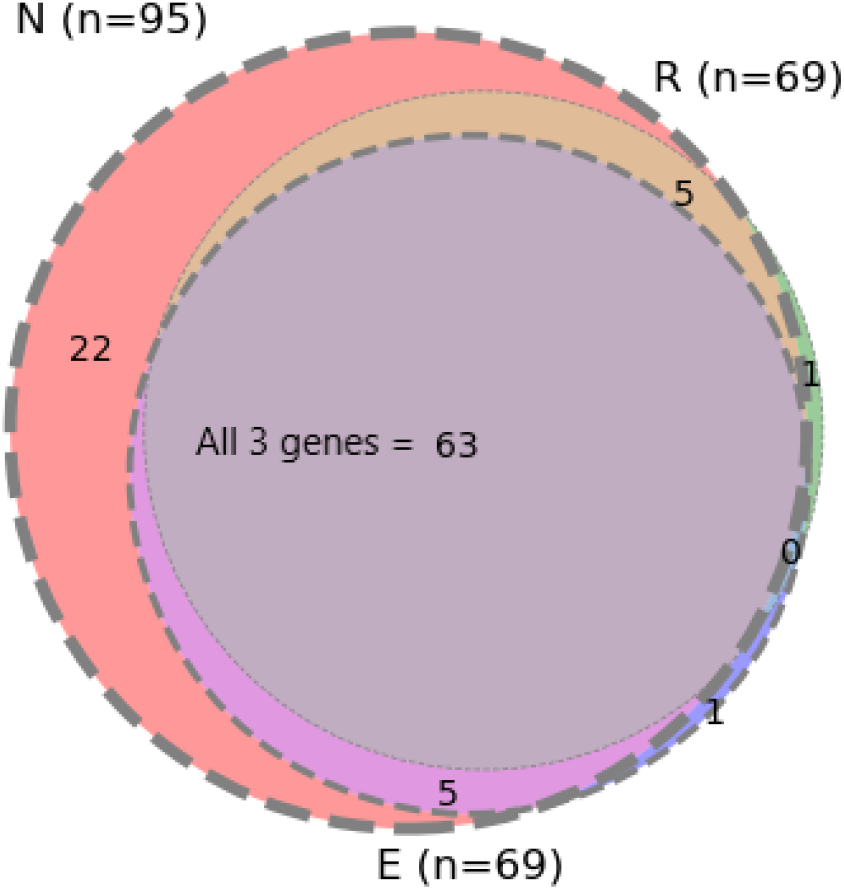
Number of positive results of **Saline Mouthwash (N=137 tests)** in each target (N, E, R genes) and their overlapping.

The median of Cq values of all positive ONP swabs (all three genes) was **30** and the average was **29.6** (SD ± 6.1), compared to a median of **32** and an average **32.2** of positive MW (SD± 4.3). While there were positive ONP swabs with negative MW in the same gene, the medians of those positive Cq values of ONP swabs, which were missed by MW were 35-36. Two MW samples were positive (Cq= 33 and 38) while the ONP swab was negative by all three genes.

Since the N gene reaction seemed to be the most sensitive marker for infection, we compared the performance of MW as tested by the N gene target in comparison to ONP swabs tested positive by each of the three targets E, R and N gene. Figure 2 shows that MW tested for the N target (n=95, out of 137 (**69.3%)** samples) detected 3.7% more samples than the number detected by R target (**65.7%**, 90 out of 137 samples) of the swab and equal to the number detected by E target.

**Figure 2:**
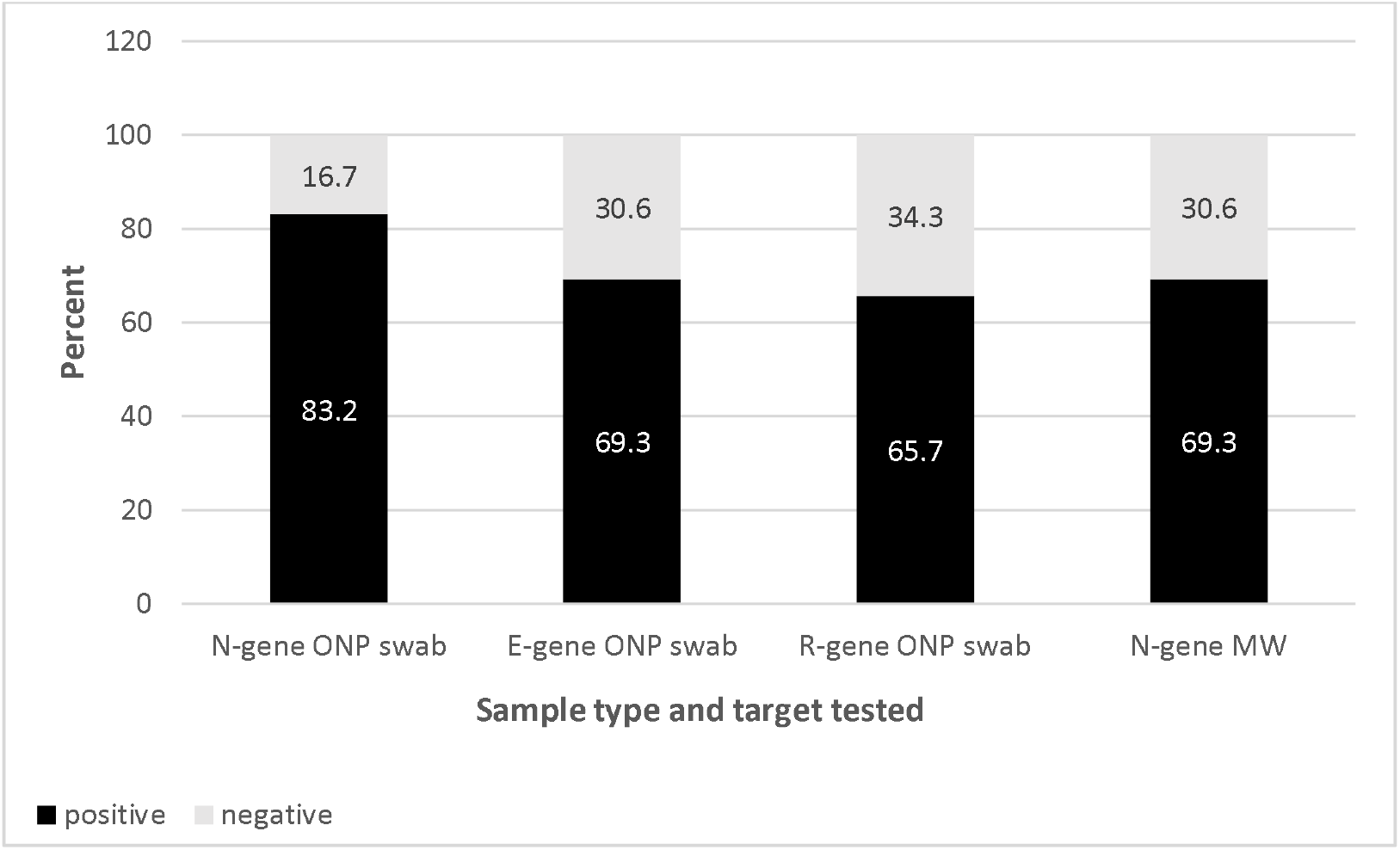
MW as tested by the N target in comparison to ONP swabs tested by N, E and R targets (n=137) **N-gene ONP swab=** positive samples of ONP swabs tested by N target; **E-gene ONP swab**= positive samples of ONP swabs tested by E target; **R-gene ONP swab**= positive samples of ONP swabs tested by R target; **N-gene MW**= positive samples of MW tested by N target

Our laboratory considers Cq values of 40 and below as positive samples, which is the reference of the above results. According to this, the MW detected **83.6%** of positive ONP swabs (table 1). When comparing the MW and ONP swabs at a different positive cutoff; Cq values of 30 and below (higher viral load), which is the accepted threshold of noninfectious state, MW as tested by the N reaction was positive (Cq ≤ 40) in detecting 94-97% of these cases.

**Table 1:** Performance of standard ONP gene targets compared to MW target N gene.

| | Positive $\leq 40$ | |
| --- | --- | --- |
|  | Positive/Total<br>Mean Cq | Missing positive cases/ compared to positive N-target<br>Mean Cq (of missed N target) |
| Swab N target | 114 /137 (83%)<br>29.9 ($\pm 5.9$) | / |
| Swab E target | 95/137 (69%)<br>28.8 ($\pm 6.5$) | 20/114 (18%)<br>36.3 ($\pm 2.0$) |
| Swab R target | 90/137 (66%)<br>30.2 ($\pm 5.9$) | 24/114 (21%)<br>36.4 ($\pm 1.9$) |
| MW -N target<br>(positive $\leq 40$) | 95/137 (69%)<br>32.9 ($\pm 4.0$) | 22/114 (19%)<br>34.6 ($\pm 3.5$) |

### Other Mouth rinse solutions

#### Distilled water

The performance of saline MW was tested in comparison to 10ml of distilled water.

Since the N gene reaction appeared to be the most sensitive marker for infection, we compared the performance of MW as tested by the N target in comparison to water wash tested by N gene. MW tested by N target detected 38 tests as positive, out of 59 tests (**64%**), equally to the detection by water.

The average of Cq values of all positive saline MW in the <u>N</u> target was **33.5** (SD ± 4.0) with a median of **35**, compared to an average of **32.9** in positive water wash in the <u>N</u> target (SD± 3.88), with a median of **34**.

### Other mouth wash solutions

A. **Listerine**®-a commercial MW containing alcohol.

Performance of Listerine MW vs saline MW was tested in 12 people who performed the paired tests. The comparison demonstrated that saline MW tested for gene N was detected in 7 out of 12 tests (**58%**), while Listerine MW tested for N target was detected in only 5 tests as positive, out of 12 tests (**42%**).

The average of Cq values of all positive saline MW in the N target was **31.1** (SD ± 5.0), with a median of 31, compared to an average of **33.6** in positive Listerine MW in the N target (SD± 3.2), with a median of 32.

B. **Orbitol**®-A commercial MW without alcohol.

We compared the performance of saline MW as tested for the N target in comparison to Orbitol MW tested for N target. The comparison shows that saline MW tested for the N target detected 13 out of 16 tests (81%) as positive, while Orbitol MW tested for the N target detected 8 out of 16 tests (50%) only.

The average of Cq values of all positive saline MW in the N target was **32.9** (SD ± 3.2) with a median of **33**, compared to an average of **36.2** in positive Orbitol MW in the N target (SD± 2.9) with a median of **36**.

Figure 3 shows the comparison of positive tests in ONP swabs as tested by the N target alone (as well as by all three gene targets) and the different MW solutions tested in this study.

**Figure 3:**
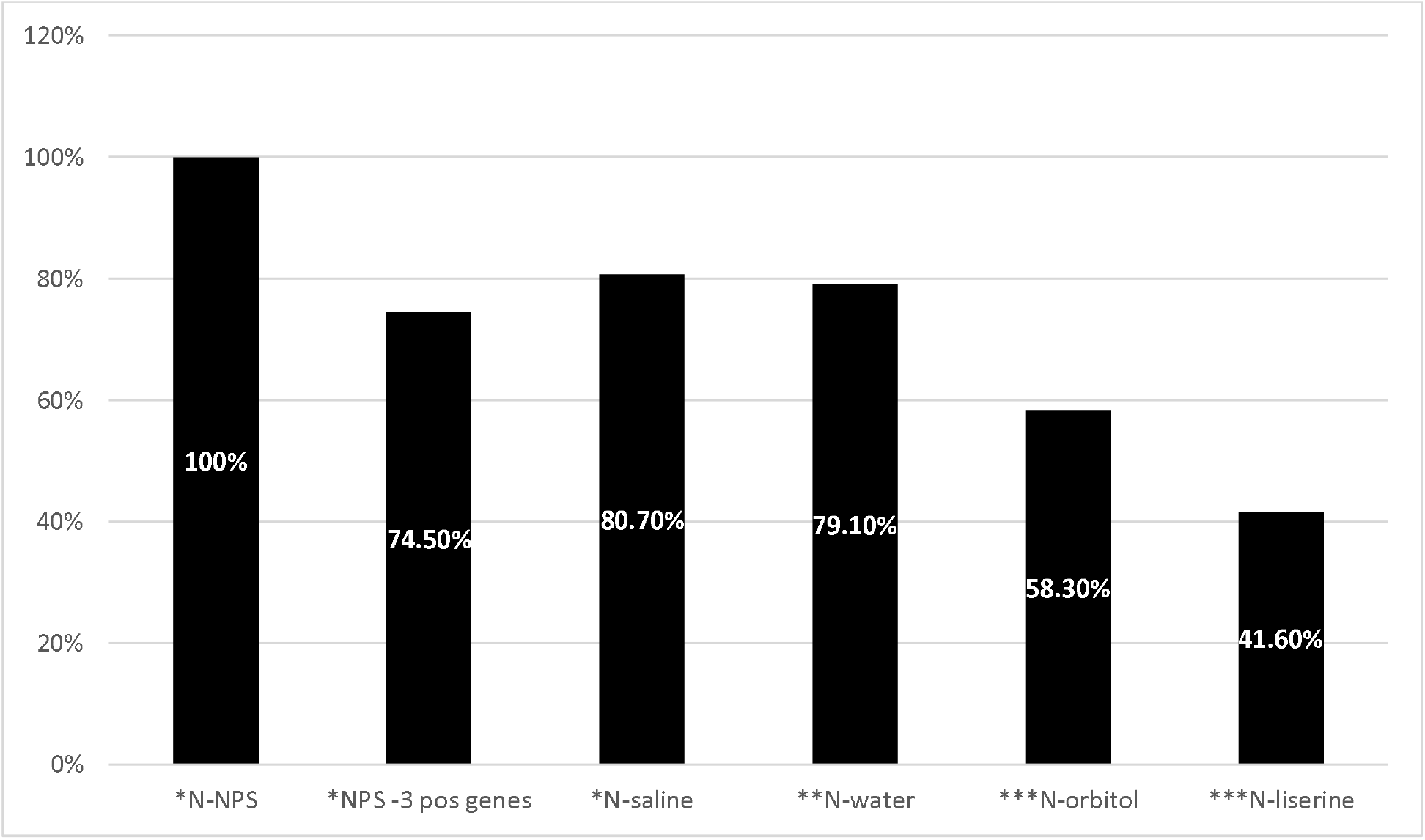
The performance of different mouth-fluid wash in comparison to positive samples by ONP swabs as tested by N-target. **N-ONP**= positive samples of ONP swabs tested by N target; **ONP-3 pos genes** = ONP swabs positive in all 3 gene targets (N,E,R) **N-saline**= positive samples of saline-MW tested by N target ; **N-water**= positive samples of water-MW tested by N target **N-Orbitol**= positive samples of Orbitol-MW tested by N target ; **N-Listerine** = positive samples of Listerine-MW tested by N target * 137 samples were tested, among them 114 were positive in ONP tested by N target (=100%). **59 samples were tested, among them 48 were positive in ONP tested by N target (=100%). ***16 samples of Orbitol were tested, among them 12 were positive in ONP tested by N target (=100%). 12 samples of Listerine were tested, among them 12 were positive in ONP tested by N target (=100%).

## Discussion

There is an extensive ongoing research aimed to establish the best sampling test for COVID-19. Thus far, the nucleic acid amplification tests (NAAT) method based on PCR amplification of respiratory sample is the gold standard. CDC guidelines have recommended the performance of swab insertion in specific areas of the respiratory tract such as the nasopharynx, oropharynx, nasal turbinate, and anterior nares.^13^ The nasopharyngeal area is still considered the preferred site. However, with its inconvenience, cost, world shortage of supplies and even safety^14^, other sampling methods are urgently needed.

In this study, we compared 137 paired saline MW and ONP swab samples, from 96 confirmed COVID-19 outpatients. Among this group of patients in different stages of their illness, including convalescence samples, RT-qPCR positivity rate was as high as 84.6% based on ONP swabs. The results of the main solution examined, MW with normal saline, showed a high detection rate - as demonstrated by high positivity rate in comparison to ONP swabs (97 positive MW out of 116 positive ONP swabs - 83.6%). Negative results were mainly at high Cq levels [average of 34.4-35], corresponding to the convalescence stage of the disease when patients are not considered as infective.^15, 16^

Our results showed a positivity rate of 83.6% MW samples in relation to positive ONP swabs upon using Cq cutoff of ≤40 (being considered as negative), yet a comparison with ONP swabs cutoff of ≤ 30 (higher viral load) yielded much better results of N-MW. It is important to mention that the new antigen rapid diagnostic tests which are commonly used, are considered less sensitive than RT-PCR. The sensitivity ranges from 0% to 94%, depending on the viral load in the specimen. However, the value of these antigen tests is the ability to detect viral load of Cq ≤30, which is considered infectious state. ^16,17,18^

Another important finding in our study was the higher sensitivity of the N gene reaction (84%), compared to E (69%) and R (65%) reactions, by ONP swabs. Different laboratories and organizations around the world use different RT-qPCR tests with different target genes, to detect SARS-2-COV. Some use a single target assay, targeting the R gene, the E gene, the Orf1ab region, or other viral genes. The clinical sensitivity varies between kits^19^, thereby potentially leading to false negative results, especially with low viral loads. The high sensitivity of N gene reaction implemented in the Seegene kit was reflected both in the ONP swabs and MW tests. The MW tested by N gene reaction (n=95; 69% positivity rate) appears to be superior to R reaction in ONP swab (n=90; 66%) and equivalent to the E reaction (n=95; 69%).

Notably, our comparison was performed where the swab sampling was based on oral and nasopharyngeal sampling, which is considered as the most sensitive approach,while other countries use different collection sites (nasopharynx, nasal turbinate, anterior nares).

Additionally, dedicated professional staff performed the procedure, thus we most likely obtained the best possible results. Moreover, the MW sampling was always performed before the ONP swab, to avoid falsely raising the amount of epithelial cells in the MW. The high positivity rate of the MW in our study (specifically the N target) may therefore offer an alternative for ONP especially in situations where there is shortage of swab sampling resources, it has no side effects compared to ONP swabs^14^, and the MW can be easily self-collected.

We compared normal saline MW with other solutions, and none were found to be superior. Water MW had an equal performance to saline MW in contrast to other solutions, which had lower detection rate. We assumed that fluids that contain lipids and alcohol (Listrine®) rinse off the epithelial cells of the oro-nasopharynx, thus producing more positive samples. However, in practice, the results showed low positivity rate, similarly to non-alcohol containing solution (Orbitol®), which resulted in low positivity rate.

In the past, studies have examined the effectiveness of gargle samples for the molecular detection of respiratory infections. Results were overall encouraging regarding the sensitivity of mouth wash compared to saliva sampling or nasopharyngeal swab, although not consistently and patient sample was very small in some of the studies (table 2).

**Table 2:** Mouthwash performance in detecting different respiratory pathogens.

| **Pathogen** | **Comparison**<br>**between**<br>**sampling**<br>**methods** | **Results** | **Reference** |
| --- | --- | --- | --- |
| Varies respiratory pathogens-The most dominant was Influenza A (18) ,then Influenza B (11) and RSV (6). | 79 NP swabs vs 79 mouth washes | Mouth washes demonstrated higher sensitivity.<br>18 positive washes with negative swabs (Cq >29), 8 positive swabs with negative washes (Cq>28). | 20 |
| COVID-19 | 24 NP swabs vs 24 mouth washes | 5 Mouth washes were positive with negative swabs. | 21 |
| Mycobacterium Tuberculosis | 127 mouth washes vs 127 sputum specimens | Mouth wash was less sensitive than sputum specimens (73% vs 99%). | 26 |
| SARS | 17 mouth wash vs 17 saliva samples | 17 Mouth washes were positive vs14 positive saliva samples. | 22 |
| SARS | 17 mouth washes | 11 Mouth washes were positive out | 27 |
|  | of serology<br>confirmed SARS<br>patients | of 17 |  |

Numerous saliva-testing studies have been conducted comparing it to different respiratory tract swab testing. Results are conflicting but there is FDA approval for some of these methods.^12^ However, saliva testing is less convenient for the patients^23^, and might be more complicated to process in the lab due to its high viscosity and presence of inhibitory saliva proteins^24^, which may further increase laboratory workload.

Comparing the different methods of obtaining respiratory tract samples is therefore a complicated task due to the high number of variables. It depends on whether a swab is taken by a medical personnel or by self-testing, it depends on the part of the respiratory tract from which the sample is obtained: nasopharynx, oropharynx, nasal turbinate, or anterior nares since there different results^25^.

The idea of self-collected specimens was examined in a study which compared nasopharyngeal swab and at least one self-collected sample - saline mouth rinse or saliva. Saline mouth rinse samples had a sensitivity of 98% (39/40) compared to 79% (26/33) of saliva samples. In addition, when patients ranked the sample acceptability (1=lowest, 5=highest), the mouth rinse had the highest mean acceptability (4.95), significantly more acceptable than healthcare worker collected NP swab (3.17) or saliva sampling (4.44).^23^

## Limitations

Our study included COVID-19 outpatients, most of them with mild symptoms, thus the study population did not reflect a population suitable for screening measures, nor did it include children. Nevertheless, there were different degrees of illness and many negative patients (in the convalescence stage) to compare the different methods among healthy subjects. The RT-qPCR assay used here included three different targets (N, E, R), which may be complicated to analyze and cannot be implemented in many laboratories. Therefore, if MW sampling will be used widely, in many cases it may not be tested by the Seegene N reaction, as our results suggest is beneficial, for sensitive detection.

On the other hand, our comparison is advantageous since the MW was compared to ONP swab (and not just NP swab), presumably the most sensitive method currently used as a standard. This sampling was performed by professional personnel, and not by the patient himself/herself, and the samples were all analyzed at the same laboratory. Under these strict conditions, we found evidence that support the effectiveness of MW as an alternative sampling method. We therefore speculate that when swabs are self-collected, or taken from only one area (nasopharynx, oropharynx, nasal turbinate, or anterior nares) there is a high probability that the MW will perform even better, in terms of sensitivity, than the swab sampling.

The results described herein suggest that using the least expensive and more convenient approach, by saline MW sampling, may be a reliable alternative for swab testing, with high positivity rate – 83.6% out of positive ONP swabs. Our finding that saline MW sampling is a reliable method of detecting SARS-2-COV, may therefore be successfully used for diagnosis of other respiratory pathogens.

## Data Availability

Data will be made available on reasonable request.

## Declarations

### Funding

The authors have declared no sources of funding.

### Conflicts of interest

The authors have declared no conflicts of interest.

### Availability of data and material

Data will be made available on reasonable request.

### Author’s contributions

D.L., A.B., O.E., E.S.: literature review, manuscript drafting. A.B, M.M, G.H, A.S, O.E: collecting data. D.L, A.B, M.M, O.E, Y.L, E.S: interpretation of data. D.L, Y.L, O.E, E,S.: critical review.

### Ethics approval

Ethical approval was obtained from Sheba Medical Center. The trial was done in accordance with the principles of the declaration of Helsinki.

### Consent to participate

Informed consent was obtained from all patients.

### Consent for publication

The manuscript has been seen and approved by all authors, who accept full responsibility for the content. The authors had full access to the data and their analysis, as well as drafting the article or editing an author’s draft.

## Notes

### Competing Interest Statement

The authors have declared no competing interest.

### Author Declarations

Ethical approval was obtained from Sheba Medical Center and informed consent was obtained from all patients .The trial was done in accordance with the principles of the declaration of Helsinki.

